# Somatosensory burst peripheral nerve stimulation focally upregulates corticospinal and spinal excitability in the upper limb

**DOI:** 10.1101/2025.05.28.25328530

**Authors:** Nish Mohith Kurukuti, Hamidollah Hassanlouei, Xin Sienna Yu, Jose L. Pons

**Affiliations:** Legs + Walking lab, Shirley Ryan Abilitylab, Chicago, USA; Department of Biomedical Engineering, McCormick School of Engineering, Northwestern University, Chicago, USA; Department of Physical Medicine and Rehabilitation, Northwestern University, Chicago, USA

**Keywords:** Transcutaneous electrical nerve stimulation, burst stimulation, corticospinal excitability, spinal excitability, motor-evoked potentials, F-wave, H-reflex

## Abstract

Peripheral nerve stimulation (PNS) is commonly used in research and clinical settings for pain management and for augmenting somatosensory input for motor recovery. Its functional effects are dependent on stimulation parameters such as frequency, intensity, and duration of stimulation. Recently, interest in temporally modulated PNS (burst PNS), in which high-frequency carrier pulses are demodulated to low-frequency bursts, has increased. Burst PNS applied below the motor threshold (sensory) has been used to suppress pain and tremor. However, the effects of burst sensory PNS (sPNS) on corticospinal and spinal excitability are unknown, limiting its application. We evaluated the impact of a session of burst sPNS on corticospinal excitability through motor-evoked potentials (MEPs) and spinal excitability through F-wave and H-reflex assessments targeting the first dorsal interosseous (FDI) and flexor carpi radialis (FCR) muscles. Ten healthy participants underwent a randomized crossover study with two experimental visits, in which corticospinal and spinal excitability were evaluated before and after a session (40 min) of burst sPNS at the wrist or no stimulation (control). Compared with the control condition, burst sPNS resulted in a focal increase in MEP amplitudes (p < 0.001) in the FDI muscle, but not in the FCR muscle (p = 0.26). Similarly, only the F-wave amplitude increased following burst sPNS (p = 0.008) for the FDI muscle compared to the control condition, but no differences were observed in the H-reflex amplitude (p = 0.33) in the FCR muscle between the burst sPNS and the control condition. Our findings suggest that burst sPNS modulates spinal and corticospinal excitability in the short term (5–10 min in this study). However, the relative changes in cortical and spinal levels due to burst sPNS are unknown, and the timeline for these continued aftereffects requires further investigation.

**Trial registration:** NCT04501133

## BACKGROUND

Peripheral nerve stimulation (PNS) is a common rehabilitation technique that is widely used to enhance motor function by modulating neuronal excitability. Clinically, it is used to alleviate chronic pain symptoms (1), enhance somatosensory input to improve motor recovery in stroke (2) and spinal cord injury (3), and suppress tremors in movement disorders (4). The PNS provides differential effects depending on stimulation parameters. The two common parameters that vary in the clinical setting are the frequency and intensity of stimulation. Motor PNS (mPNS), which is delivered above the motor threshold to evoke extra torque/force, has been used to augment motor rehabilitation and/or assist in generating functional movements (gait) in individuals with stroke and incomplete spinal cord injury (5, 6). Alternatively, sensory PNS (sPNS), with a stimulation intensity below the sensory threshold when delivered at high frequencies (> 90 Hz), is used to suppress pain (1) and tremor (4), whereas at low frequencies (< 30 Hz) in combination with target-oriented training, it is used to rehabilitate hand function in stroke (7).

Mechanistically, the PNS evokes artificial volleys that propagate orthodromically and antidromically along recruited afferent and/or efferent fibers. Orthodromic afferent volleys travel to the spinal cord and brain modulating corticospinal (CS) excitability. This modulation persists for up to 2 h after stimulation (8). Studies using transcranial magnetic stimulation (TMS) have shown that the modulation of motor evoked potentials (MEPs), a measure of CS excitability, is dependent on PNS parameters such as frequency (9, 10), intensity (11), duration (12), and even the pattern or waveform of delivery (13). Recently, interest in temporally patterned sPNS to improve its effectiveness in rehabilitation has increased. Burst sPNS, in which high-frequency carrier pulses are demodulated to low-frequency bursts, has shown promise in suppressing symptoms such as pain (14) and tremors (15). However, the impact of such a burst-modulated sPNS on CS and spinal excitability remains largely unexplored, despite evidence suggesting that the pattern of stimulation delivery plays a critical role in neuroplasticity.

Emerging data suggest that the temporal structure of afferent inputs, not just the total pulse count or intensity, can shape neurophysiological outcomes. In a recent study, Ishibashi et al. (13) reported that intermittent (burst-like) sPNS increased MEP amplitude, an indicator of enhanced corticospinal excitability, whereas continuous sPNS at the same frequency and duration decreased excitability. This underscores the unique neuroplastic potential of the burst-patterned input, potentially owing to its resemblance to natural afferent firing patterns and its ability to prevent synaptic fatigue. Understanding how this specific burst sPNS protocol affects CS and spinal excitability could have significant clinical implications, potentially enhancing rehabilitation strategies.

The modulation of MEP amplitudes has been largely attributed to changes in cortical excitability (16). Following short-term (1–2 h) high-frequency PNS, the somatosensory cortical representation of the muscles innervated by the stimulated nerve increases (17). Similar increases in corticomotor representation in the motor cortex following high-frequency PNS have been reported. Imaging studies have also shown that the PNS induces cortical and subcortical excitability changes in sensorimotor regions (18). These findings highlight the importance of proprioceptive afferent engagement in modulating the sensorimotor circuits. However, the involvement of spinal mechanisms in the modulation of CS excitability remains unclear. While studies using H-reflex and F-waves have shown no changes in spinal excitability following PNS (11, 13), recent findings suggest an increase in spinal excitability following high-frequency PNS when delivered with wide pulse widths (19). These inconsistencies regarding the effects of the PNS on spinal excitability highlight the need for further research.

In this study, we investigated the effect of burst sPNS using a wide pulse width targeting the median, radial, and ulnar nerves at the wrist and compared it to that of a control condition (no stimulation) by examining the CS and spinal excitability in the first dorsal interosseous (FDI) and flexor carpi radialis (FCR) muscles. We hypothesized that MEPs are evoked by TMS over the motor cortex, and that the F-wave amplitude evoked by ulnar nerve stimulation increases after burst sPNS in the FDI. We also hypothesized that an increase in corticospinal and spinal excitability would only be present in the FDI but not in the FCR muscle because of the focal effect of burst sPNS (20). By evaluating these expected changes, we aimed to contribute to the growing body of knowledge on how afferent volleys generated during burst sPNS alter CS and spinal excitability in humans, which could inform the development of more effective rehabilitation protocols.

## METHODS

### Participants

Ten healthy adults (mean age: 33.7 ± 13.0 years; 5 females) volunteered to participate in this randomized crossover study. The participants did not have a history of neuromuscular disorders, sensory deficits, or upper limb surgery. Written informed consent was obtained from all participants prior to testing. This study was approved by the Northwestern University IRB Review Board (IRB: STU00211930), and all methods conformed to the standards of the Declaration of Helsinki (2004). All subjects participated in two separate ∼3-h testing visits at least 72 h apart, in which burst sPNS was applied to the median, radial, and ulnar nerves at one visit and no stimulation (control) at the other visit. The order of visits was randomized for each participant by block randomization. The random allocation sequence based on participant IDs was generated using R. The time of day of each session was the same for each participant to reduce the potential confounding effect of diurnal changes in CNS excitability (21). The subjects were instructed to avoid caffeine consumption 12 h prior to the testing sessions and during a session to eliminate its influence on CNS excitability (22) and to avoid intense physical activity 12 h prior to the testing sessions. Following the first visit, participants had a 3-day washout period before participating in the second visit.

### Motor-evoked potentials and recruitment curves

Motor-evoked potentials (MEPs) were recorded from the FDI and FCR muscles via surface electrodes on the dominant arm. The EMG signal was pre-amplified and digitized at 2048 Hz. Relaxation was defined as EMG activity at baseline of < 20 µV peak-to-peak amplitude for at least 1 s. During the setup, the optimal scalp positions (contralateral motor cortex to the dominant hand) to elicit reliable MEPs (at least five out of 10 attempts) for the FDI and FCR muscles were mapped. For the remainder of the visit, TMS was delivered to the optimal scalp position to stimulate the FDI and FCR muscles. TMS was delivered through a figure-eight shaped magnetic coil (outer diameter of 8.7 cm) connected to a MagPro X100 stimulator (MagVenture, Denmark). The magnetic coil was placed tangentially to the scalp, with the intersection of both wings at a 45° angle with the midline, to optimally stimulate the motor cortex (Brasil-Neto et al. 1992; Mills et al. 1992). The stimulation location was marked on a TMS cap secured on the participant’s head to ensure the repeatability of coil placement throughout the experiment.

In addition to mapping the optimal scalp position, the resting motor threshold (rMT), defined as the minimum TMS intensity (measured to the nearest 1% of the maximum output of the magnetic stimulator) required to elicit at least five of ten MEPs ≥ 50 µV in consecutive trials (23), was also determined for the FDI and FCR muscles. For the remainder of the visit, TMS was delivered at intensities expressed relative to the rMT measured in the muscles. The mean MEP amplitudes were obtained in response to 10 TMS stimuli delivered at each of the six stimulus intensities: 90%, 100%, 110%, 120%, 130%, and 140% of the rMT for each muscle, with the order of intensities randomized.

### Maximal M-waves (M-max)

To determine the M-max for the FDI and FCR muscles, the stimulation intensity was increased over several stimuli from below the motor threshold to 1.5–2 times the minimum current required to evoke M-max (20). The M-max was calculated as the largest M-wave evoked in the muscles of the three trials. The amplitude of M-max from each muscle was tested before and after delivery of the burst sPNS or in the control condition.

### F-waves

F-waves were evoked to examine motor neuron excitability in the FDI muscle using supramaximal stimulus intensity to the ulnar nerve (200-μs pulse duration; DS5; Digitimer). Sixty stimuli were delivered at 1 Hz, with an intensity of 150% of M-max (24). For each stimulus, the peak-to-peak amplitude and persistence (i.e., the percentage of stimuli evoking a response) of F-waves were measured. F-wave trials were filtered via a second-order Bessel high-pass filter (200 Hz) to “flatten the tail of the M-wave” (25). An F wave was considered present if a response with a proper latency (minimum of 30 ms after the stimulus artifact) had an amplitude of ≥ 20 µV.

### H-reflex

H-reflexes were elicited to examine spinal excitability of the FCR muscle by stimulating the median nerve (1-ms pulse duration; DS5; Digitimer) near the elbow. The response was identified as an H-reflex if it had a latency between 12 and 25 ms. The intensity to elicit H-max was determined by systematically increasing the intensity and quantifying the H-reflex and M-wave amplitudes (H-M curve). The intensity at which the amplitude of the H-reflex was the maximum with the minimal M-wave was considered the maximum intensity (25). Ten stimuli were delivered approximately 5 s apart at H-max intensity to record the H-reflex at H-max. The peak-to-peak amplitude of the H-reflex was measured for each stimulus.

### Burst sPNS

During the experimental session with burst sPNS, electrical stimuli were delivered to nerves at the wrist. A bipolar constant current stimulator (Digitimer DS5, Digitimer, UK) was used along with conductive surface electrodes (2 cm diameter; Axelgaard, Denmark) to deliver the electrical current. Two anodes were placed over the median nerve at the flexor retinaculum on the palmar side, and over the radial nerve at the distal radius of the wrist. A common cathode is placed over the distal end of the ulna. Burst sPNS involves trains of rectangular biphasic wave pulses with a duration of 800 µs at a carrier frequency of 100 Hz, which were applied at 5 Hz (100 ms on and then switched off for 100 ms) and delivered for 40 min. The stimulus intensity was adjusted to avoid visible contraction of the hand muscles while providing paresthesia, but not being painful or uncomfortable. This low-intensity stimulation and stimulus duration preferentially activates large cutaneous and proprioceptive sensory fibers (26). The participants were instructed to refrain from moving their stimulated arm during the administration of the burst sPNS.

### Data analysis

Changes in CS excitability induced by burst sPNS were determined by quantifying and comparing the group averages of 10 MEPs evoked before and after burst sPNS or the control condition. To ensure that all MEPs were obtained at rest, MEP data were inspected post hoc and discarded if the EMG during the 1 s before TMS exceeded two standard deviations of the average baseline signal recorded at rest before stimulation. Of the 2,350 MEPs evoked by 10 subjects, 19 MEPs (∼1% of the total responses) were removed from the analyses based on this criterion. To compare the change in CS excitability, the average MEP amplitude from 10 trials in post-assessments for each muscle was normalized to the average MEP amplitude in the pre-assessments using Equation (1).

Changes in spinal excitability induced by burst sPNS were determined by quantifying and comparing the average peak-to-peak amplitude of the F-waves evoked in the 60 trials and the H-reflex from 10 trials before and after the burst sPNS or control condition. The percentage occurrence of F-waves was also computed before and after the burst sPNS and control conditions. To compare the changes in spinal excitability, the percentage changes in the average peak-to-peak amplitudes of the F-wave, H-reflex, and F-wave persistence were computed using Equation (1).

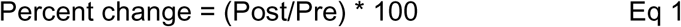

### Statistical analysis

All statistical analyses were performed using R software. The normality of data distribution was checked using quantile_quantile plots and histograms. Analyses were performed using linear mixed effect models implemented in the *lme4* function with the Kenward-Roger method to estimate the denominator degrees of freedom and p-values. This method considers and accounts for the dependence of the data points within each participant. When necessary, multiple comparisons were performed using the package *emmeans*, which adjusts the p-value for multiple comparisons using the Tukey method. The significance level was set at p = 0.05. The values are reported as mean ± standard deviation.

To evaluate the effect of burst sPNS on CS excitability across muscles, percent changes in MEPs for FDI and FCR were compared using linear mixed-effect models with muscle (FDI, FCR) and condition (control, burst sPNS) as fixed effects and participants as random effects. To evaluate the effect of burst sPNS on spinal excitability, we compared the percent change in F-wave amplitude, percent change in H-reflex amplitude, and percent change in F-wave persistence via linear mixed-effect models with condition (control, burst sPNS) as a fixed effect and participants as a random effect. To examine the effect of burst sPNS on the M-max for each muscle, we also compared the M-max amplitudes via a linear mixed-effect model with condition (control, burst sPNS) and time (Pre, Post) as fixed effects and participants as random effects.

## RESULTS

### Maximal M-waves (M-max)

The maximal M-wave amplitudes for the FCR muscle did not significantly affect time (F = 0.42, p = 0.52) or condition (F = 0.042, p =0.83). Similarly, the maximal M-wave amplitudes for the FDI muscle did not significantly affect time (F = 0.45, p =0.51) or condition (F = 2.50, p =0.18).

These findings suggest that Mmax did not differ due to burst sPNS or the control conditions for both the FDI and FCR muscles.

### Motor evoked potentials

Significant effects of condition (F = 10.38, p = 0.002), muscle (F = 16.37, p < 0.001), and the interaction between condition and muscle (F = 38.02, p < 0.001) were observed for the percent change in MEP amplitudes for the FDI and FCR muscles. Post hoc analyses revealed that the percent change in MEPs increased for the FDI muscle due to burst sPNS compared with the control condition (137.4 ± 39.8% for burst sPNS vs 97.6 ± 15.9% for the control; p < 0.001; Fig 2D). However, there was no difference in MEPs for the FCR muscle due to burst sPNS compared with the control condition (94.2 ± 21.6% for burst sPNS vs 106.5 ± 30.8% for the control; p = 0.26; Fig 2E). This suggests that burst sPNS had a focal increase in MEP amplitude in the FDI muscle, but not in the FCR muscle. Furthermore, the increase in MEP amplitudes following burst sPNS suggests an increase in corticospinal excitability in the FDI due to burst sPNS.

**Figure 1:**
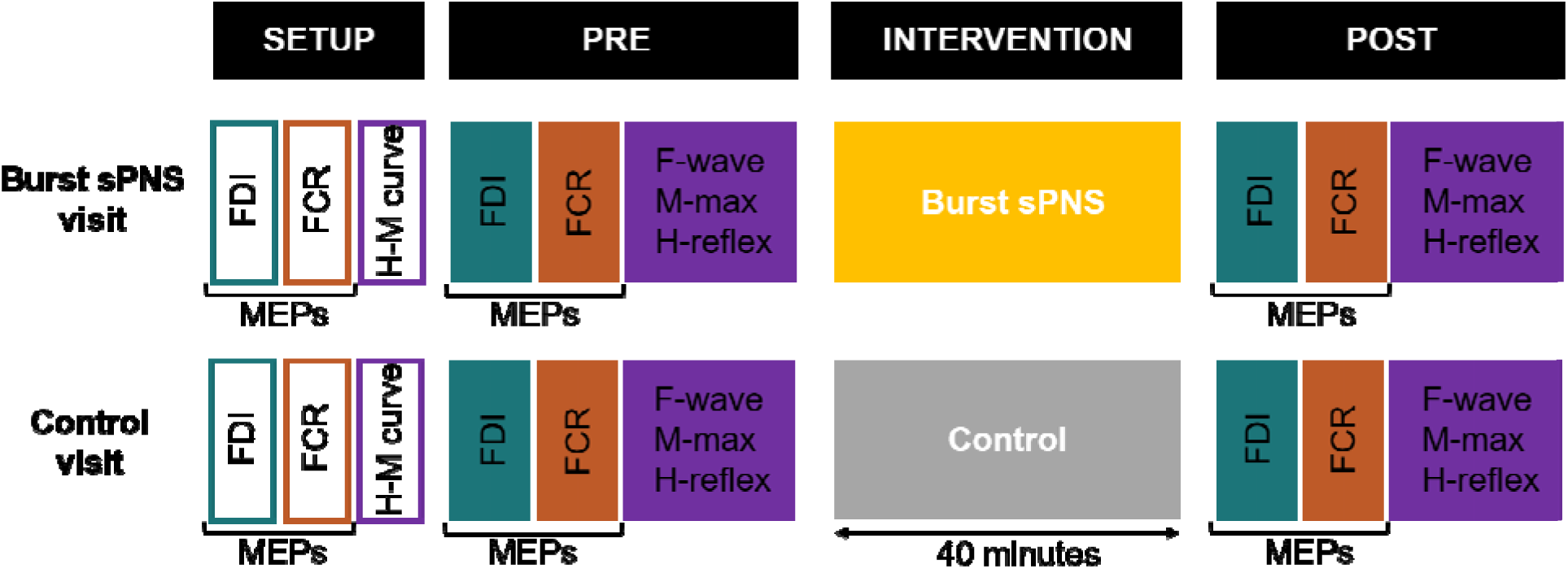
Experimental paradigm. The participants underwent two experimental visits, one with burst sPNS and the other with control stimulation. The order of experimental visits was randomized across participants. In the setup session, the cortical hotspots for eliciting motor-evoked potentials (MEPs) and the resting motor thresholds for the FDI and FCR muscles were determined. The Hreflex-Mwave curve was also mapped during the setup session to determine the H-max and M-max for the FCR and FDI muscles. Electrophysiological outcomes such as the MEP input□output curve (FDI and FCR), H-reflex (FCR), F-wave (FDI), and M-max (FDI and FCR) were measured before (Pre) and after (Post) the intervention session (burst sPNS or control). The intervention session lasted for 40 minutes.

**Figure 2:**
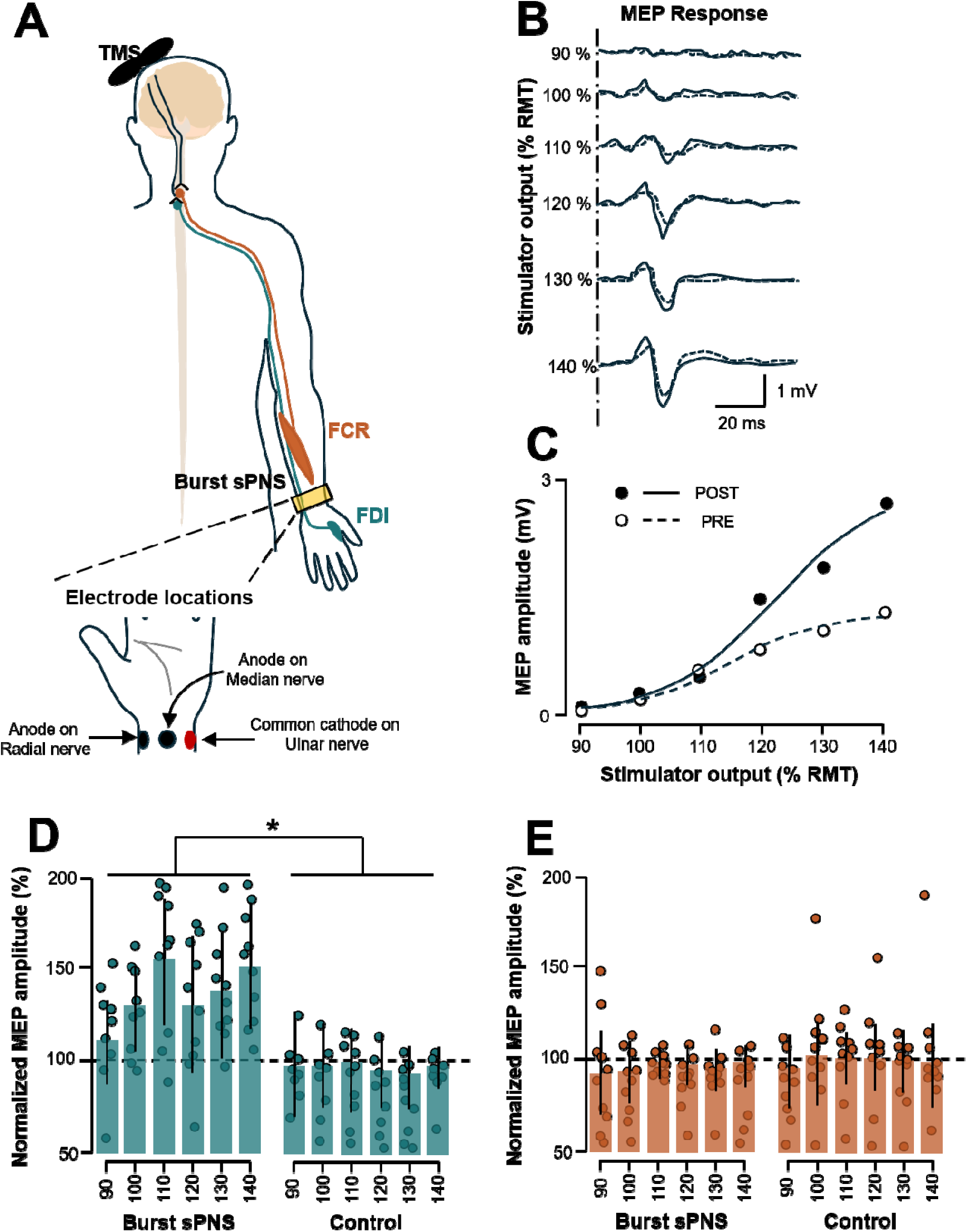
Corticospinal excitability. (A) MEPs were evoked in the FDI and FCR muscles in the dominant hand via TMS targeting the hotspots for the respective muscles from the contralateral cerebral cortex. Once the resting motor threshold (RMT) was determined for each muscle (FDI and FCR), six intensities from 90% - 140% of the RMT were recorded before and after the bust sPNS or control condition. The burst sPNS was delivered to the nerve at the wrist, with the anodes on the radial and median nerves and a common cathode on the ulnar nerve. The peak-to-peak amplitude was quantified to compute the MEPs at each intensity to map the input□output curve of the MEP. An example of a participant’s MEP responses at various intensities tested and the input□output curve is shown in B and C. Changes in MEP amplitude were quantified via the average MEP amplitude from the PRE and POST sessions to evaluate the impact of burst sPNS compared with the control (no stimulation) condition. The change in MEP amplitude increased for the FDI muscle (D) following burst sPNS compared with the control condition across all intensities. However, no differences were observed in the MEP amplitudes for the FCR muscle (E) due to burst sPNS. The scale bars shown represent the means, and the error bars indicate SDs; *P□<□0.05.

### F-waves

A significant effect of condition (F = 12.7, p = 0.008) was observed for the percent change in F-wave amplitudes for the FDI muscle. Post hoc analysis revealed an increase in the F-wave amplitude following burst sPNS compared to the control condition (120.1 ± 17.1 for burst sPNS vs 103.2 ± 16.84 for control; p = 0.008). However, no significant effect of condition (F = 2.74, p = 0.13) was observed for the percent change in F-wave persistence for the FDI muscle (Fig 3). This suggests that burst sPNS increased motoneuronal excitability, but did not affect the persistence of the F wave between the two conditions.

**Figure 3:**
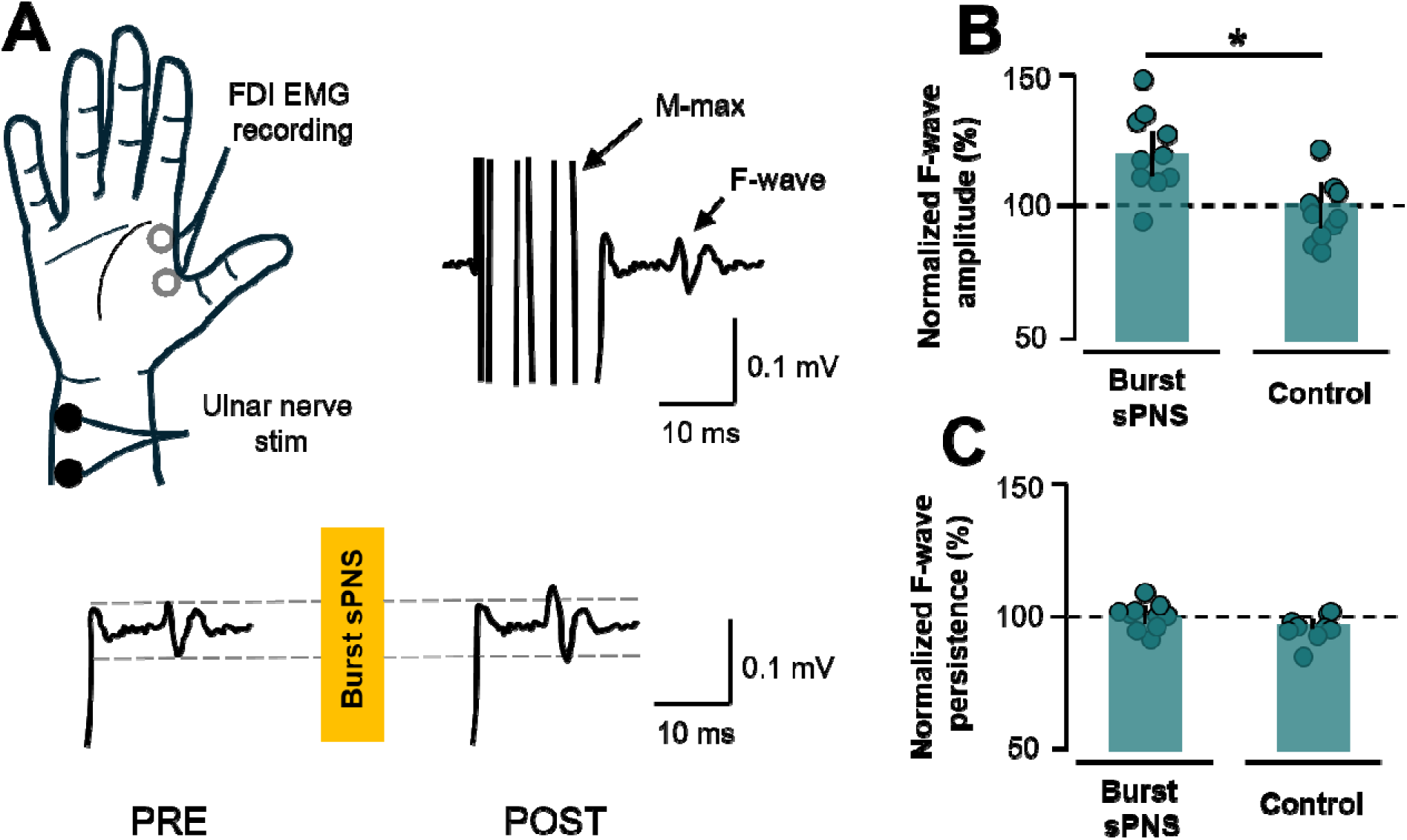
Spinal excitability at the FDI muscle. Spinal excitability at the FDI muscle was measured through F-waves by stimulating the ulnar nerve at 150% of the M-max (A). An example of the change in the F-wave due to burst sPNS is illustrated (A, bottom panel). Changes in F-wave amplitude (B) and F-wave persistence (the ratio of trials that evoked F-waves to total trials, C) were quantified across the interventions. The percentage change in F-wave amplitude increased due to burst sPNS compared with the control condition, but no difference was observed in F-wave persistence. The scale bars shown represent the means, and the error bars indicate SDs; *P□<□0.05.

**Figure 4:**
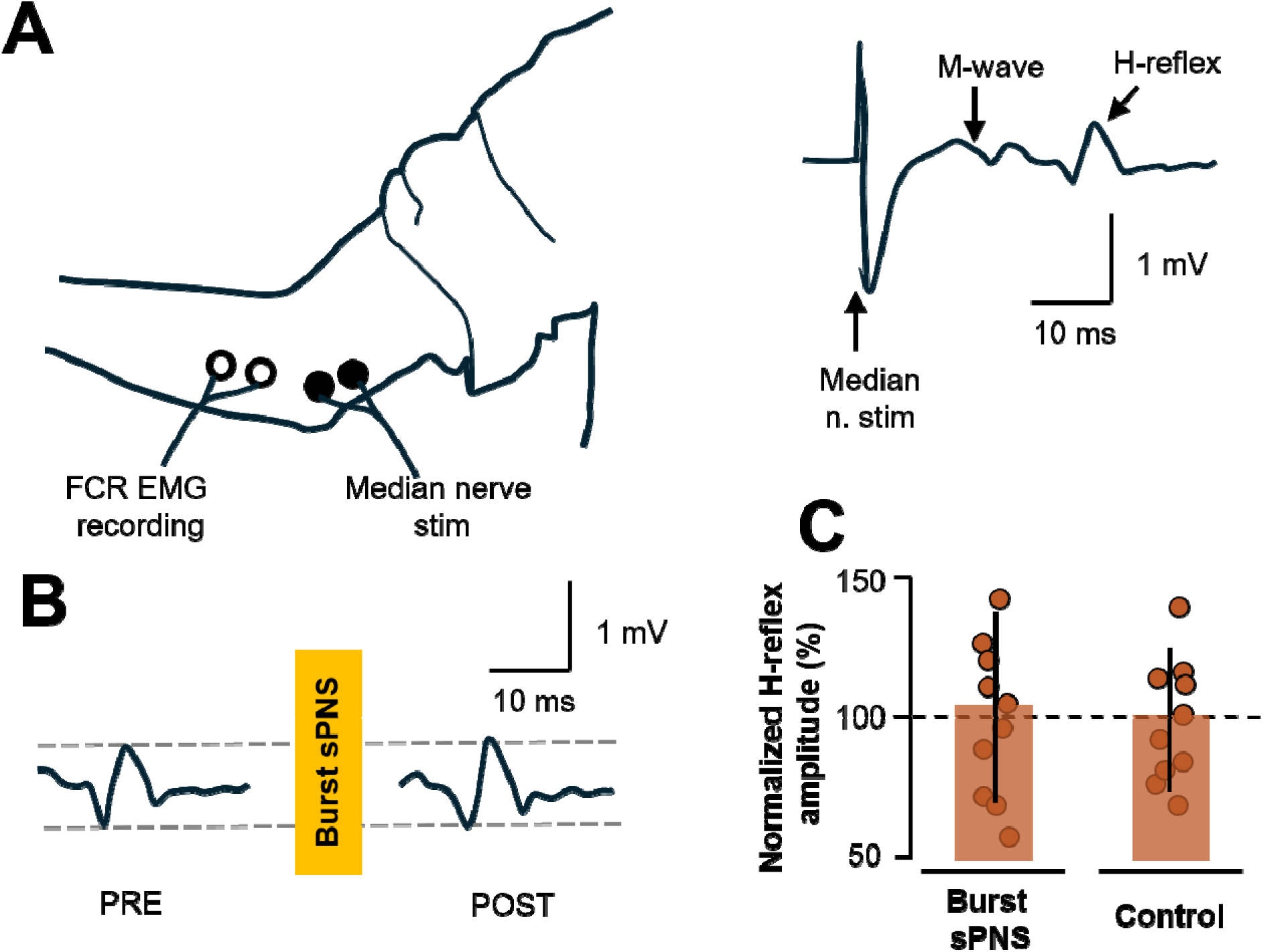
Spinal excitability at the FCR muscle. H-reflexes were evoked from the FCR muscle by stimulating the median nerve near the elbow (A). An example of a change in the H-reflex due to burst sPNS is illustrated (B). The percentage change in the H-reflex did not significantly differ between the burst sPNS condition and the control condition (C). The scale bars shown represent the means, and the error bars indicate the SDs.

### H-reflex

No significant effect of condition (F = 1.15, p = 0.332) was observed for the percent change in H-reflex amplitudes for the FCR muscle, suggesting that burst sPNS did not modulate the spinal excitability of the FCR muscle (Fig 3).

## DISCUSSION

The aim of the present study was to examine the effects of burst sPNS on corticospinal and spinal excitability and whether these effects were focused on the muscle innervated by the stimulated nerve. Our findings revealed that burst sPNS uniquely increased corticospinal and spinal excitability in the muscle distal to the stimulation site but had no significant effect on the muscle proximal to it. Specifically, we observed that MEPs and F-wave amplitudes increased for the FDI (distal muscle), but neither MEPs nor H-reflex amplitudes were modulated in the FCR (proximal muscle) following burst sPNS. In this study, we demonstrated that burst-modulated subthreshold stimulation can increase MEPs and F-waves, suggesting that burst delivery may mimic or amplify proprioceptive engagement typically associated with contraction, possibly by synchronizing the afferent volleys and modulating corticospinal and spinal excitability in the targeted muscle.

It is well documented that the PNS, which is an agnostic stimulation parameter, modulates CS excitability in muscles innervated by the stimulated nerve in the upper limb (20, 27–29). Our findings corroborate the hypothesis of region-specific effects of PNS. The differences observed between the FDI and FCR muscles indicate that afferent volleys elicited by burst sPNS predominantly modulate the neural pathways of the muscles distal to the site of the stimulated nerves, underscoring its potential for precise motor rehabilitation. However, to our knowledge, this is the first study to show that burst sPNS, where bursts of high-frequency stimulation are delivered, increases corticospinal and spinal excitability in the target muscle.

### Corticospinal excitability with burst sPNS

Several studies have demonstrated that PNS modulates CS excitability (18). This modulation has been attributed to changes in the somatosensory cortex and, consequently, in the motor cortex due to the direct somatotopically organized corticocortical connections between the somatosensory and motor cortices (18). Similarly, the observed increase in corticospinal excitability in the FDI muscle following burst sPNS can be attributed to repetitive activation of afferent pathways from the ulnar nerve-innervated muscles distal to the stimulation site, producing convergent input to the sensorimotor cortex. Furthermore, the findings from this study further extend this knowledge by showing that burst timing, even in the absence of overt muscle contraction, can facilitate excitability, echoing the findings of Ishibashi et al. (13). reported that intermittent (burst) PNS increased MEPs, whereas continuous PNS of equivalent intensity and frequency decreased MEPs. This finding suggests that the temporal structure of afferent input plays a critical role in facilitating plasticity mechanisms, potentially through Hebbian-like processes, where afferent input strengthens corticospinal connections specific to FDI. In support of this, previous studies in animal models (17) and humans (30, 31) have demonstrated the role of afferent stimulation in reorganizing cortical circuits and increasing motor cortical excitability.

Functional imaging studies corroborate our findings, showing that sPNS enhances cortical excitability in both healthy individuals (31) and in stroke patients with corticospinal tract damage (32). sPNS increases the signal intensity and number of activated voxels in the primary motor cortex (M1) during motor tasks, as observed through blood oxygen level-dependent (BOLD)-based fMRI. Additionally, arterial spin labeling revealed increased perfusion in the M1 at rest following sPNS in healthy individuals, likely driven by increased neuronal activity and associated blood flow (33). Together, these findings reinforce the hypothesis that burst sPNS promotes plastic changes in sensorimotor cortical circuits, underscoring the ability of burst sPNS to improve motor function.

### Spinal excitability with burst sPNS

Our observation of increased F-wave amplitude without changes in the M-max or H-reflex amplitude supports the interpretation that burst sPNS enhances spinal motoneuron excitability, particularly at the postsynaptic level. In our study, M-max did not change due to burst sPNS, which is consistent with previous findings (11, 13). Many previous studies using tonic or continuous PNS protocols failed to show spinal changes (10, 34). Studies that used intermittent mPNS (11, 13), where twitches were observed in the target muscle during stimulation (above motor threshold intensity), were stimulated with a 30 Hz frequency and a narrow pulse width of 0.2 ms. In the present study, we used a higher carrier frequency (100 Hz), which was demodulated to a much lower frequency (5 Hz), delivered at an intensity set below the motor threshold, with a wide pulse width of 0.8 ms. Similar to our PNS protocol, Vitry et al. used a wide pulse (1 ms) stimulation at 100 Hz and demonstrated increases in thoracic-evoked MEPs (19). A narrow pulse width has been shown to predominantly facilitate direct peripheral motor recruitment, with less contribution through central mechanisms. In contrast, a wider pulse width preferentially engages proprioceptive and large-diameter cutaneous afferents, which may more effectively influence central circuits and evoke motor recruitment through relatively greater central pathways (35). This could explain the disparity in the spinal excitability results.

Modulation of spinal excitability at rest can be achieved through either presynaptic inhibitory mechanisms or changes in the intrinsic properties of motoneurons (36–38). An increase in spinal excitability of the FDI muscle can include compensatory mechanisms at the presynaptic and/or postsynaptic levels that compensate for the inhibitory effect of repetitive afferent stimulation. At the presynaptic level, a potential reduction in the sensitivity of Ia afferents to presynaptic inhibition is proposed as a possible mechanism. In anesthetized cats, repetitive high-frequency activation of Ia afferents leads to increased calcium accumulation in Ia terminals and, consequently, an increased probability of neurotransmitter release (39), which lasts up to several minutes following sustained high-frequency stimulation (40). At the postsynaptic level, changes in intrinsic motor neuron properties can explain increased spinal excitability at rest, as observed following burst sPNS. Persistent inward current (PIC) activation is a plausible mechanism as it is sensitive to neuromodulatory inputs originating from the brainstem’s caudal raphe nucleus and locus coeruleus (41). These brainstem regions respond to electrical stimulation applied to a nerve (42), suggesting that the volleys from the burst sPNS may reach the brainstem and regulate the descending neuromodulatory input. When the raphe spinal pathway is stimulated, synaptic release of serotonin on dendritic and somatic 5-HT2 receptors promotes Ca+2 PICs (43). Hence, these presynaptic and/or postsynaptic mechanisms may contribute to the changes in spinal excitability.

### Clinical and mechanistic implications

Together, these findings support the use of burst sPNS as a targeted, noninvasive strategy for priming the corticospinal and spinal circuits. In contrast to conventional TENS or continuous sPNS, burst-modulated protocols offer a temporally structured afferent signal that may better mimic natural sensory inputs and avoid habituation. Similarly, previous studies by Jadidi et al. (44) reported that burst and PWM-modulated TENS enhanced motor cortical excitability, suggesting that patterned stimulation protocols may induce more robust and spatially selective plasticity than traditional methods. Given the focal effects observed in the FDI muscle and the lack of change in the FCR muscle, our results highlight the spatial specificity of burst sPNS, which may be a valuable characteristic for applications in stroke, spinal cord injury, and movement disorders, where regional modulation is essential. Furthermore, the ability to increase excitability without requiring volitional effort or overt contractions opens doors for early intervention in severely impaired individuals.

### Future work

Earlier investigations utilized a diverse range of stimulation parameters, including variations in the intensity, frequency, pulse widths, patterns, and electrode placement. While many studies have focused predominantly on either spinal or corticospinal circuits [for example, (34, 44)], recent investigations have evaluated both corticospinal and spinal circuits together, emphasizing the need for deeper exploration of these mechanisms (19, 45). Although burst sPNS enhances both corticospinal and spinal excitability through potentially distinct yet complementary mechanisms, the relative contributions of these pathways cannot be distinguished from the current experimental design. Future studies should be structured to specifically isolate and quantify the contribution of each site to the resulting increase in motor output. A clear understanding of these contributions could inform the development of targeted rehabilitation strategies for neurological disorders involving dysfunction of the corticospinal or spinal pathways. Longitudinal studies are needed to determine whether these acute excitability changes translate into sustained functional improvements when paired with training. Additionally, optimizing the burst parameters (e.g., carrier frequency, burst duration, and duty cycle) is crucial for clinical translation.

## CONCLUSION

Our findings underscore the potential of burst sPNS as a non-invasive and adaptable tool for enhancing motor recovery in neurological disorders. By increasing both corticospinal and spinal excitability, burst sPNS could serve as a “priming” technique and may support functional improvements when combined with physical therapy, personalized neuromodulation, or diagnostic applications. This innovative approach not only offers a promising avenue for motor rehabilitation but also paves the way for its application in performance enhancement and broader neuromodulatory strategies.

## Data Availability

Raw data were generated at the Legs + Walking Lab of Shirley Ryan Abilitylab. The derived data supporting the findings of this study are available from the corresponding author (JLP) upon request

## DECLARATIONS

### Ethics approval and consent to participate

Written informed consent was obtained from all the participants before testing. This study was approved by the Northwestern University IRB Review Board (IRB: STU00211930), and all methods conformed to the standards of the Declaration of Helsinki (2004).

### Consent for publication

Not applicable

### Authors’ contributions

Conceptualization: NMK and JLP; Data curation: NMK, HH, XSY; Formal analysis: NMK, HH; Funding acquisition: JLP; Investigation: NMK, HH, XSY, JLP; Methodology: NMK, HH; Project administration: NMK, JLP; Resources: JLP; Software: NMK; Supervision: JLP; Validation: NMK, HH; Visualization: NMK, HH; Writing-original draft: NMK, HH; Writing – review and editing: NMK, HH, XSY, JLP.

## Acknowledgments

The authors would like to thank Grace W. Hoo for helping with subject enrollment and study management.

